# Curating, collecting, and cataloguing global COVID-19 datasets for the aim of predicting personalized risk

**DOI:** 10.1101/2021.11.14.21265797

**Authors:** Sepehr Golriz Khatami, Maria Francesca Russo, Daniel Domingo-Fernández, Andrea Zaliani, Sarah Mubeen, Yojana Gadiya, Astghik Sargsyan, Reagon Karki, Stephan Gebel, Ram Kumar Ruppa Surulinathan, Vanessa Lage-Rupprecht, Saulius Archipovas, Geltrude Mingrone, Marc Jacobs, Carsten Claussen, Martin Hofmann-Apitius, Alpha Tom Kodamullil, the COPERIMOplus consortium

**Author notes:** **Corresponding Authors:** Tom Kodamullil, A., Department of Bioinformatics, Fraunhofer Institute for Algorithms and Scientific Computing (SCAI), Sankt Augustin 53757, Germany.

## Abstract

The COVID-19 data catalogue is a repository that provides a landscape view of COVID-19 studies and datasets as a putative source to enable researchers to develop personalized COVID-19 predictive risk models. The COVID-19 data catalogue currently contains over 400 studies and their relevant information collected from a wide range of global sources such as global initiatives, clinical trial repositories, publications and data repositories. Further, the curated content stored in this data catalogue is complemented by a web application, providing visualizations of these studies, including their references, relevant information such as measured variables, and the geographical locations of where these studies were performed. This resource is one of the first to capture, organize and store studies, datasets and metadata in the area of COVID-19 in a comprehensive repository. We are convinced that our work will facilitate future research and development of personalized predictive risk models of COVID-19.

## 1. Introduction

Coronavirus disease 2019 (COVID-19) has spread globally and become a public health emergency. As of July 2021, approximately 210 million cases have been reported in which around 4.5 million people have died^1^. Moreover, COVID-19 has deepened economic suffering and set an unprecedented economic recession globally. Although the rapid vaccination campaign has led to a reduction in the number of cases in the countries with a high vaccination rate, a deeper understanding of the disease to reduce its burden is needed.

COVID-19 is a potentially fatal respiratory illness that predominantly affects the lungs, but can also damage several other organs, such as the heart, brain, liver, and kidneys **[1]**. A wide spectrum of research studies have analyzed COVID-19 related risk factors **[4,5,6]** and treatments **[7,8,9]**. While most people recover from COVID-19 without needing hospital treatment, COVID-19 can be fatal in older individuals (≥60 years) as well as persons with certain medical conditions (e.g., diabetes, hypertension, obesity, etc.), often requiring hospital-based treatment such as oxygen supplementation for these individuals **[2,3]**. Nonetheless, fatal progression of disease in younger patients as well as in re-infected patients is also being observed **[10]** and the human response to COVID-19 infection appears to be very heterogeneous **[11]**. This heterogeneity among infected individuals calls for modelling approaches that can predict the risk of a given patient with the ultimate goal of preventing and personalizing their treatment.

Data-driven personalized models have the potential to predict individualized risks as they can identify more relevant and informative patient-specific risk factors based on previously generated patient-level data **[12]**. However, there are several factors that need to be considered about the pre-existing data in order to build personalized predictive models, such as adequate sample size, relevant endpoints, the types of variables, and the longitudinal aspect of datapoints **[13]**. Experimental results show that with increasing training data, the accuracy and robustness of data-driven models can be improved, which in turn could lead to better predictions and the use of these models in day-to-day clinical practice **[14,15]**. Thus, the first step in building personalized COVID-19 predictive models is to generate a detailed overview on the available trial and study data suited for the training of COVID-19 personalized models.

From the start of the COVID-19 pandemic, various data catalogues (i.e., services or tools that manage various datasets in a central metadata directory) have been established. Some of them are publicly accessible are openICPSR (https://www.openicpsr.org/openicpsr/search/studies), the Microsoft Azure COVID-19 data catalogue (https://azure.microsoft.com/de-de/services/open-datasets/catalog/bing-covid-19-data), the Amazon COVID-19 data catalogue (https://aws.amazon.com/data-exchange/covid-19), and the World Bank data catalogue (https://datacatalog.worldbank.org).While, the studies listed in the aforementioned data catalogues focus on general information such as hospital capacities, participant mobility, and social distances, they ignore relevant features that are critical for building predictive models (e.g., biomarkers). Moreover, the number of collected studies in these data catalogues is limited (<100 datasets) as, for example, the Microsoft Azure COVID-19 data catalogue has 35 studies while the World Bank data catalogue has 29. For these reasons, although the organization of this information and published data are critical for predicting and better understanding COVID-19 epidemiology, they are not suitable for training and developing personalized predictive models as they lack longitudinal datapoints and clinical variables from hospitalized patients.

To address these shortcomings, we collected and curated COVID-19 datasets from clinical trials, publications, and other global initiatives, building a comprehensive COVID-19 data catalogue as a source to bring together published findings within the context of study information, study design and other related variables which could inform risk modelling approaches. We collected information from more than 400 studies which not only include general study information, such as number of patients, study location and patient outcomes, but also contain more specific information for each individual study including measurement class (e.g., intensive unit care (ICU), clinical, laboratory), the measured features of a particular class (e.g., oxygen saturation, lymphocyte), and study type (e.g., observational, interventional).

The resulting COVID-19 data catalogue provides a comprehensive and interactive view of results from an extensive, systematic search and curation effort of several hundreds of global initiatives and clinical trials registries, and scientific articles. We prioritized the collected studies based on different criteria including, ***i)*** number of patients, ***ii)*** data availability, ***iii)*** measured variables (e.g., ICU, clinical, laboratory), ***iv)*** one-time measurements (cross-sectional data) or multiple-time measurements (longitudinal data). Next, based on the study rank, the corresponding principal investigators (PIs) were contacted to acquire the datasets of interest. Since the data structure and format of a study are defined based on the study’s goal and the available resources, they are not compatible with each other, nor are they directly usable for downstream applications. Thus, in the last step, the acquired datasets were harmonized, normalized, and curated to be applicable for machines to develop and train COVID-19 personalized predictive models. Finally, the presented resource lays the groundwork for modelling approaches that support the identification of patient-specific risk factors which can ultimately improve our understanding of the disease.

## 2. Materials and Methods

The major objective of this work was to generate a comprehensive data catalogue which contains the data and variables necessary to model individual progression of COVID-19. To reach our goal, we designed a workflow with five main steps, namely: 1) study selection, 2) data extraction and quality assessment, 3) data acquisition, 4) data curation, harmonization and variable mapping, and 5) dataset preparation (**Figure 1**). The corresponding activities of each step are described in sections 2.1 to 2.5.

**Figure 1.**
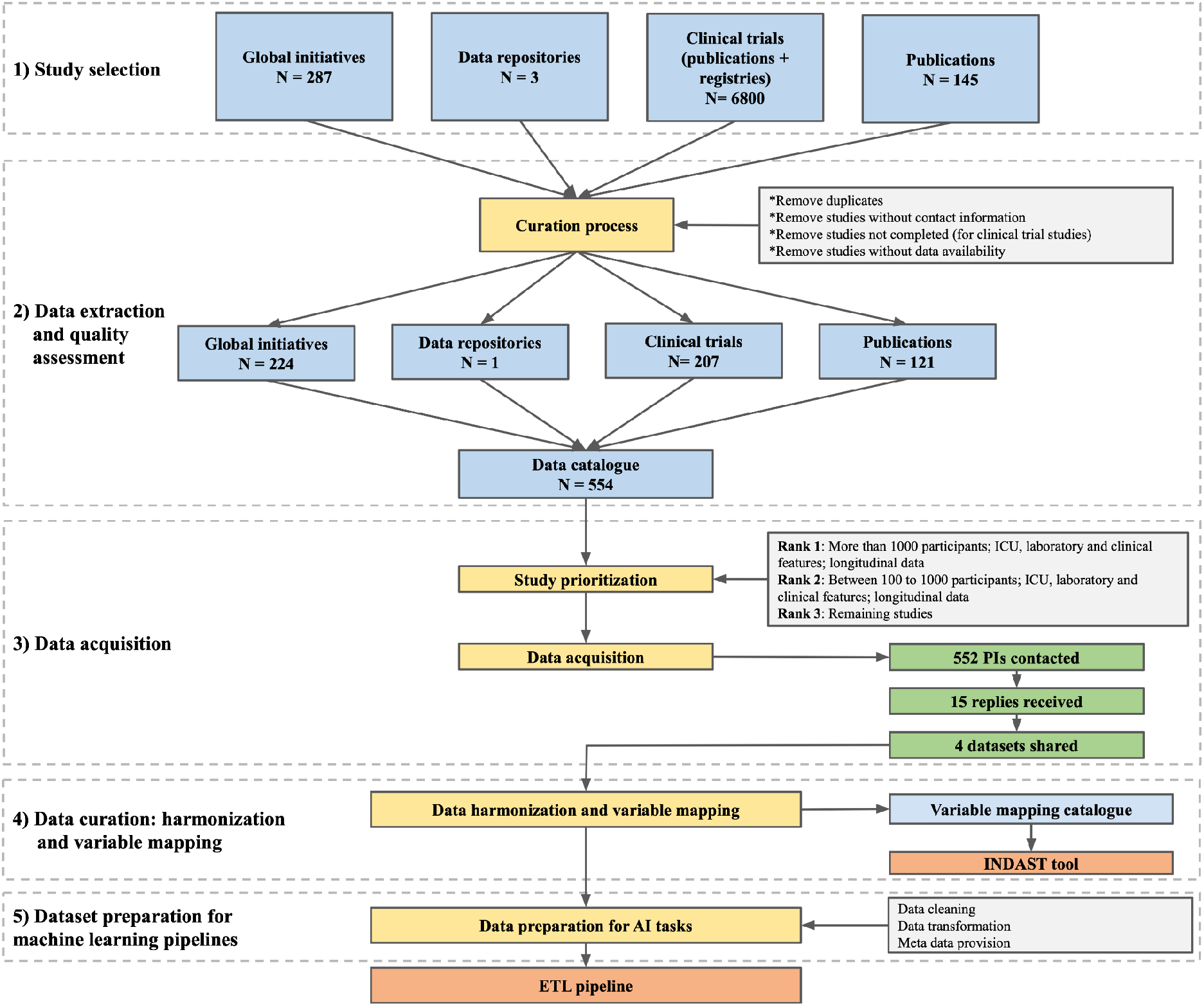
Overview of the steps required for the preparation of the data catalogue. The workflow includes five major steps from study selection to the preparation of datasets for machine learning pipelines. Detailed explanations of each step are described in the corresponding subsections below.

### 2.1. Study selection

Studies included in our data catalogue were identified via four different routes: global initiatives, clinical trial repositories, publications and data repositories.

#### 2.1.1. Global initiatives

These encompass i) COVID-Analytics (https://covidanalytics.io/dataset), a group of researchers from the MIT Operations Research Center, ii) COVID-NMA (https://covid-nma.com/), iii) COVID-evidence (https://covid-evidence.org/database), a continuously updated database of worldwide evidence on interventions for COVID-19, iv) the COVID-19 Data Catalogue (https://datacatalog.worldbank.org/story/covid-19-data), v) the Global Health Drug Discovery Institute (GHDDI) (http://www.ghddi.org/), a transformative drug discovery and translational platform, and vi) CORONAWHY (https://www.coronawhy.org/), a globally distributed, volunteer-powered research organization attempting to assist the medical community in answering key questions related to COVID-19.

#### 2.1.2. Clinical trials repositories

Clinical trial repositories include Clinical Trials Map (HeiGIT) (https://apps.heigit.org/covid-19-map-of-hope/clinical_trials.html), Chinese Clinical Trial Registry (ChiCTR) (http://www.chictr.org.cn/abouten.aspx), and the WHO-International Clinical Trials Registry Platform (ICTRP) (https://www.who.int/clinical-trials-registry-platform), a meta-repository of registered trial information from more than 20 registries worldwide. The ICTRP provided structured information for 4,957 COVID-19 related trials (retrieved on 14.08.2020) including the number of participants, contact address, intervention and study type. However, not all of the trials are relevant for us as we need those which provide longitudinal data on COVID-19 progression for at least more than 100 participants. Based on these minimum criteria, we classified the trials into highly relevant, relevant, less relevant, and not relevant, of which only the first three trial types were added to our data catalogue. The classes and their corresponding criteria are summarized below. The complete list of criteria as well as different outcome measures are provided in **Supplementary Table 1**.

Highly relevant: Studies with at least 2 outcome measures, high participant rates, interventional studies, studies with longitudinal data and those with our outcome of interest (e.g., studies containing data of ICU or respiratory intermediate care unit (RICU) participants).

Relevant: Studies with 1 or 2 outcome measures or with a target outcome of medium interest (e.g., identification of COVID-19, mortality).

Less relevant: Studies with highly specific subject groups (e.g., pregnant women, cancer patients, healthcare workers).

Not relevant: Studies with no follow-up, very short time frames, cross-sectional studies and studies with no target outcomes.

We would like to highlight that for the studies in which two or more criteria existed, the ranking was done based on the combination of the criteria. Moreover, for the relevant studies that were identified, information of the (expected) completeness of the study and the data of contact persons (typically the PI) were also collected.

As the ICTRP platform does not contain all details required to select relevant studies (e.g., contact data, outcome measure, and study status) and is seldom updated after first registration, information was also collected from the individual registries included in the ICTRP. For example, in order to identify clinical trials described in the ChiCTR (http://www.chictr.org.cn), we first identified all studies with a Chinese clinical trial repository ID (ChiCTR followed by 10 digits) described in the ICTRP as of 27.11.2020 (https://www.who.int/clinical-trials-registry-platform), which resulted in 789 COVID-19 related clinical trials. The ChiCTR was used to obtain additional information for criteria missing from the ICTRP, such as secondary outcomes. The following steps were applied to narrow the list of studies from 789 studies (selected based on relevant criteria):

1. Nearly all studies which specified the different participant subgroups provided the number of subjects in each of them (e.g., intervention group: 80; control group: 80) and we considered the total number of participants as the sum of participants across all subgroups. Thus, we were able to select a subset of 288 studies out of a total of 789 with at least 100 or more participants, to satisfy the criteria described above.
2. Studies cancelled/retracted for any reason (e.g., due to a lack of participants) were removed from the list of target studies (i.e., 47 studies removed in total).
3. Sequential, cross-sectional and dose comparison studies were excluded, as were studies with a factorial study design.
4. The remaining clinical trials (233) were checked and ranked in terms of relevance as per the criteria described above.

Of the studies obtained from the ChiCTR, 32 studies were noted as highly relevant for AI-based risk modelling as of 01.12.2021. One limitation of the ChiCTR was the absence of information on study completeness. To overcome this limitation, trial identifiers were cross-checked in EUROPE PMC and PubMed; if publications on these trials could be found, we considered the studies to have intermediate results and have taken them into consideration. The publications retrieved also contained review articles discussing several trials, including the trial originally searched. These articles were removed and only the ones directly describing a particular study were considered as trial publications. In many cases, results may be published at a later date or may not be published at all. In addition, although longitudinal trials may not yet be completed, they may contain valuable intermediate results.

By comparing the trials from the ICTRP with the ones selected earlier (i.e., based on sources used in previously described efforts, such as our text mining approach), we noted that 25 CHiCTR trials were already included in the data catalogue, of which 13 would have been excluded when applying our above-mentioned criteria (i.e., either the number of subjects was below 100, or the study design was sequential), three were marked as highly relevant and 6 as relevant.

Additionally, we have collected publications that refer to COVID-19 clinical trial information. This more comprehensive search for publications on completed clinical trials was done by our in-house text mining tool, SCAIView, using the clinical trial identifier tagger with a text mining approach. We have generated a clinical trial identifier tagger that enabled us to search for all clinical trial IDs, e.g., clinicaltrials.gov: NCT + 8 numbers (e.g., NCT04299152) or Chinese clinical trial registry: ChiCTR + 10 numbers. Using this clinical trial identifier tagger, we have searched in the CORD19 corpus and Medline 2020. As a result, we obtained 12,247 publications that refer to 4,540 studies from 17 different clinical trial repositories. Several papers referring to multiple studies, and 4,442 publications in CORD19 containing literature on clinical trials from before 2020 were removed. Additional search criteria, such as “clinical outcome”, were also added in SCAIView. Please see the Supplementary file TM21.09

Overall, we have manually inspected each of these trials and selected 298 studies that could potentially be used for AI application based on the number of participants (more than 100), completion date (or intermediate results available before 09.2021), and study type (e.g., longitudinal).

#### 2.1.3. Publications

From the start of the pandemic, a wide spectrum of studies which focus on different aspects, such as COVID-19 related risk factors **[16]** and its treatment **[9]**, have been published. However, we aim to gather studies that are useful in developing predictive risk models in which the collection of longitudinal data is one of their main characteristics. Thus, to narrow down the publications search domain, we defined longitudinal data as one of our main keywords. In addition, in order to minimize false positive hits, we used ‘risk modelling’ and ‘prediction analysis’ as other keywords. Taken together, we applied the search paradigms ‘longitudinal data’, ‘prediction analysis’, ‘risk modelling’, ‘COVID-19’ and ‘SARS-CoV-2’ to PubMed in order to identify work that has been done in the context of COVID-19 risk prediction modelling.

In parallel to the collection of studies with any of the above-mentioned keywords, we also searched for publications in PubMed applying the search paradigms ‘serology’ and ‘serological profiling’ to specifically identify COVID-19 related articles with serological information. The goal of also collecting serological surveys and the extraction of their metadata was because this data is important for evaluating relevant factors such as the extent and duration of immunity to SARS-CoV-2 infection or possible cross-immunity. Furthermore, serological data is required to determine seroprevalence and to detect previously undetected asymptomatic infections. Another pertinent aspect is the investigation of serological details such as the study design, laboratory methods and outcome measures in relation to demographic factors in order to predict the risk of infection and the severity of the disease for different populations in society.

#### 2.1.4. Data Repositories

Studies included in our data catalogue were also identified through major data repositories, including Lean European Open Survey on SARS-CoV-2 infected patients (LEOSS), BBMRI-ERIC and European Clinical Research Infrastructure Network (ECRIN).

1. LEOSS (https://leoss.net/) is a clinical patient registry for patients infected with SARS-CoV-2 established by the joint effort from the EITaF and the German Society for Infectious Diseases. It comprises patient data from the LEOSS cohort, including patients on whom invasive ventilation was performed, and variables such as age, gender, month of diagnosis, outcome, and the presence of administered vasopressor agents.
2. BBMRI-ERIC (https://www.bbmri-eric.eu/) is a European research infrastructure for biobanking integrating information and associated samples from over 60 organizations in 20 countries to store all types of human biological samples, such as blood, tissue, cells or DNA.
3. ECRIN (https://ecrin.org/) is a non-profit organisation that regulates multinational clinical trials by linking scientific partners and networks across Europe. In general, studies which evaluated different measurement categories such as ICU, clinical, and laboratory in a longitudinal manner were included in the data catalogue. However, some studies with cross sectional data may also have been included considering the outcome of the study.

### 2.2. Data extraction and quality assessment

The metadata information contained in the resulting data catalogue were manually extracted by ten independent curators and added to the data catalogue template. The spectrum of curated metadata covers numerous fields, including study reference and title, study location, number of subjects, study type, data type, study duration, whether a variable category exists, and the measured variables for a particular category (the full list of metadata is available at https://github.com/SCAI-BIO/covid-data-catalogue). The data catalogue template was predefined based on the initial set of 15 studies. However, new fields were subsequently added to accommodate the information extracted from additional studies. Furthermore, we conducted a quality control step where entries were reviewed by multiple curators to exclusively retain those studies which fulfilled our criteria including data availability, number of subjects, phase of study (completed studies in the context of clinical trials), contact information availability, and useful data in the context of the measured variables and the number of follow-up measurements.

### 2.3. Data acquisition

The studies included in the data catalogue were divided into 3 priority levels based on various criteria, including ***i)*** the number of patients, ***ii)*** data availability, ***iii)*** measured variables (e.g., ICU, clinical, laboratory), and ***iv)*** number of follow-up measurements. Studies with longitudinal data, a substantial number of patients (e.g., 1,000 or more), variable types including ICU, clinical, and laboratory features, and data availability were assigned the highest priority level. Similarly, studies with longitudinal data, patients within a range of one hundred to one thousand, variable types such as ICU, clinical, and laboratory features, and data availability were assigned the second highest priority level, while the remaining studies were considered as the third priority. Then, based on the study rank, the corresponding PI(s) of the studies with highest priority were contacted in the first round and the other two priorities were contacted in the second and third round, respectively.

### 2.4. Data curation, harmonization and variable mapping

From the start of COVID-19 pandemic, numerous regional, national and international studies have been performed, each of which provides important insights into the disease. To develop a robust personalized predictive risk model, these studies should be semantically interoperable and the data points used by these datasets need to be normalized. However, a global standard data model that comprises all COVID-19 related terms is not available as of now. Therefore, we have used three controlled vocabularies to establish the interoperability and mapping towards various datasets and data points, namely, German Corona Consensus Dataset (GECCO) **[18]**, the COVID-19 Ontology **[19]** and the Corona Virus Vocabulary (COVOC) (https://github.com/EBISPOT/covoc/raw/master/covoc.owl).

However, assessing this interoperability is not straightforward as the studies investigated neither followed a common naming system for variables nor represented values of the same measurement in an equal manner. For example, variable names in individual datasets may be in different languages based on the origin of the study. Moreover, units of measurements in different datasets may also differ. For example, while hemoglobin may be measured based on mole per volume in one dataset, it may be measured based on gram per deciliter in another dataset. In addition, even if an overlap does exist between two variables, they may not be the same. For instance, while in a dataset the age of a sample might be explicitly mentioned, in another dataset it is implicit in the date of birth and thus must be inferred to enable data interoperability.

To enable a meaningful and comparable assessment of data interoperability, harmonization and variable mappings have been done in two steps:

1. **Variable name normalization:** All non-English variable names were translated into English using common translators. Next, a native speaker checked whether the original variable names and their translations were correct.
2. **Variable mapping:** Mapping of variables was done in a semi-automatic way. First, the variables of datasets were mapped using the *tidystringdist* string-matching package in the R programming language. The *tidystringdist* package calculates the distance between variables (all against all) of datasets (pairwise) in the range of zero (completely similar) and 1 (completely different). Next, we filtered the variables whose distance was more than 0.25. This threshold was selected after manual investigation of the variable mapping results of multiple datasets. We found that the proportion of correct mapped variables with a distance less than 0.25 was the highest among all other tested thresholds. Then, the mapped variables were manually investigated by a trained curator to check whether they were indicating the same concept (flagged 1, otherwise 0). Finally, the number of mapped variables which were flagged as 1 were considered as an interoperability score of two datasets.

After the harmonizing and variable mapping procedure, the mapped and unmapped variables were added to our variable mapping catalogue. Using this catalogue, we extended the ontology that reuses the provided GECCOplus data model and dataset (https://art-decor.org/art-decor/decor-project--covid19f). Using this ontology, we enable data stewards or data scientists to map variables from new clinical datasets to concepts in our data model via the web-based INDAST Tool (http://coperimo.mevis.fraunhofer.de/). The main goal of the INDAST Tool is to provide a platform for sharing and harmonizing new clinical datasets (not only in the COVID-19 context) as input for machine learning pipelines.

### 2.5. Dataset preparation for machine learning pipelines

The ability to build a generalizable, (personalized) predictive machine learning model based on given data is dependent on a number of criteria **(Table 1)**. In this table, we define several important criteria and assess the AI-readiness of acquired datasets based on them.

**Table 1.** AI-readiness criteria. This table summarizes the criteria which are used to evaluate the AI-readiness of the dataset.

| Dataset | Criteria | Quality |  |  |
| --- | --- | --- | --- | --- |
|  |  | Low | Medium | High |
|  | Number of patients | - | - | - |
|  | Completeness of data | - | - | - |
|  | Number of features in relation to number of patients | - | - | - |
|  | Event-rate (i.e, the fraction of patients with complete longitudinal follow-up and thus, recorded events) | - | - | - |
|  | Objectiveness of the clinical outcome (i.e., is clinical outcome a subjective or physician-dependent variation?) | - | - | - |

For the general support of downstream AI tasks and dataset AI-readiness evaluation, we performed basic data cleaning and preprocessing and compiled a data dictionary. See details in the **Supplementary File**.

### 2.6. Visualization and COVID-19 data viewer

We have developed an interface to facilitate access to the curated metadata of the datasets included in the COVID-19 data catalogue. This interface summarizes the most important metadata, such as measured features, and provides a means to obtain access to the datasets. The web application was implemented in Python using Flask and several JavaScript libraries for visualization purposes including D3.js, DataTables, and DataMaps.

## 3. Results

### 3.1. Data catalogue content

The data catalogue includes information on 553 studies which were extracted from different sources:

#### Global initiatives

In total, 287 studies were collected from the global initiatives’ sources (COVID-Analytics: 202, COVID-NMA: 64, COVID-Evidence: 21) in which after curation (i.e., omitting duplicates and studies without contact information), 224 studies were added to our data catalogue **(Figure 2)**.

**Figure 2.**
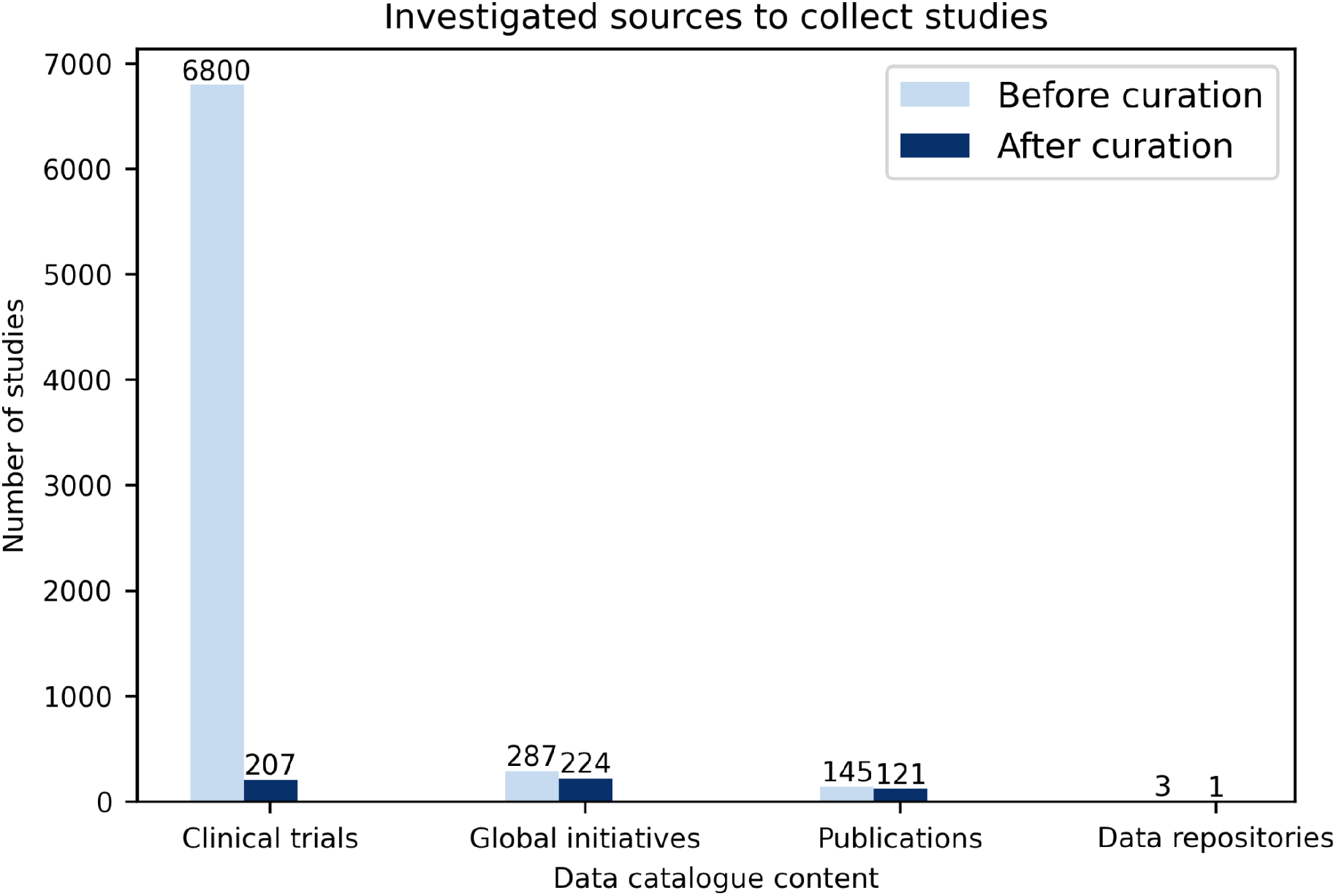
Number of studies from different sources included in the data catalogue. Three main sources including global initiatives, clinical trials and publications were investigated. Over 7,000 studies were investigated, from which, following curation, 553 studies were added to the data catalogue.

#### Clinical trials

Until 27.11.2020, around 6,800 clinical trials were registered in different registers worldwide e.g., ClinicalTrials.gov: 3,327, ChiCTR: 762, CTRI: 514, IRCT: 337 **(Figure 2)**. After extracting information from all these clinical trials such as the number of participants, contact address, study type (e.g., interventional, observational), study phase (e.g., recruiting, completed), and data type (cross sectional, longitudinal), 207 trials were selected to add to our data catalogue. The chosen trials were selected based on multiple criteria including: ***i)*** a minimum of 100 participants, ***ii)*** the availability of clinical/ICU data, ***iii)*** the availability of longitudinal data and ***iv)*** if the studies were completed.

#### Publications

Altogether, 145 studies were gathered from PubMed, bioRxiv and medRxiv using different combinations of SARS-CoV-2, COVID-19, machine learning, survival analysis, longitudinal data, and clinical data keywords. Out of the 145 studies, 121 remaining after curation were added to our data catalogue (i.e., omitting duplicate studies, those with missing contact information, and studies with explicit mentions that the data cannot be shared) **(Figure 2)**.

### 3.2. Studies overview

Not all studies included in the data catalogue addressed the same study goal, thus, the same variables were not consistently observed across these studies. For example, while some studies were designed to answer specific questions on interventions and include drugs as important variables, other studies investigated COVID-19 risk factors and thus, other types of variables, such as demographics and comorbidities were considered in them. **Figure 3** displays different types of variables which have been measured in the studies included in the data catalogue. While clinical and laboratory variable types are the two most commonly measured variables, imaging variables are the least common.

**Figure 3.**
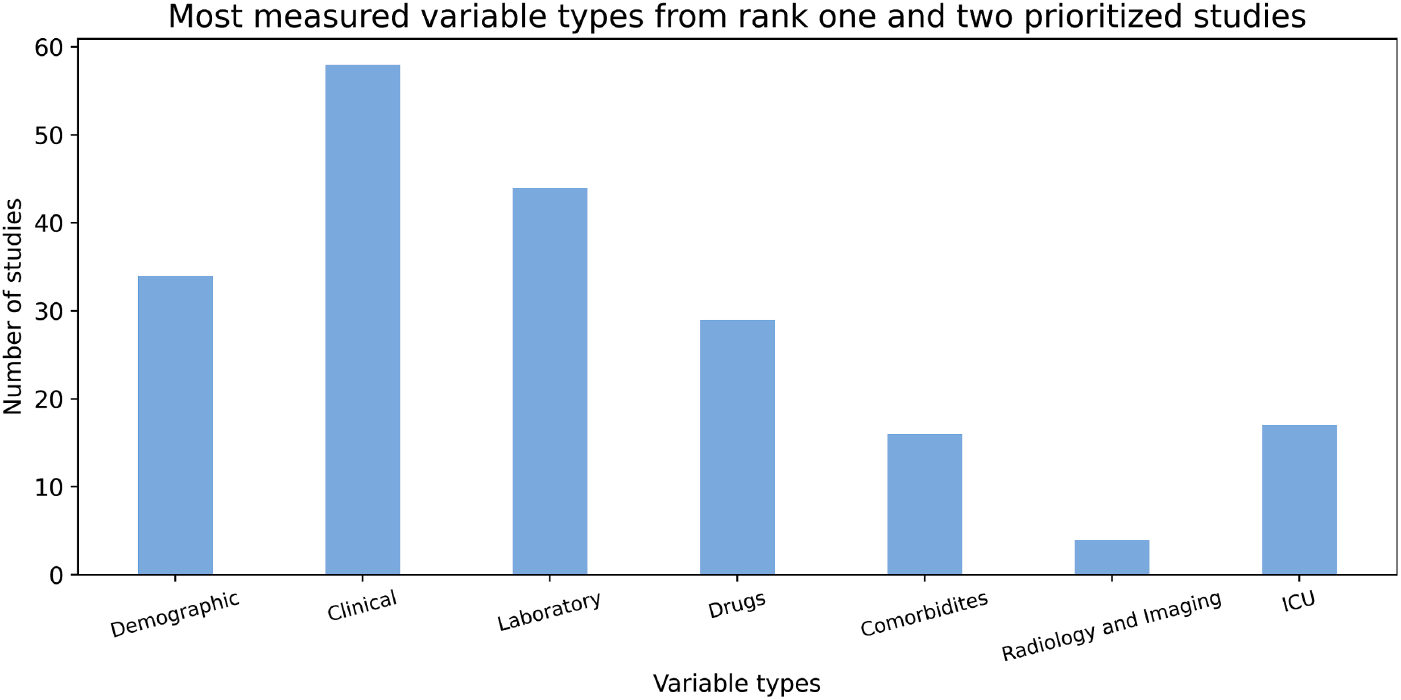
Distribution of different variable types from over 50 rank one and two prioritized studies. The clinical and laboratory variables are the two most measured features from over 150 studies.

Similarly, several associated COVID-19 clinical and laboratory variables such as blood oxygen saturation, respiratory rate, neutrophils, lymphocytes, platelets and D-dimer have been reported **[20,21]**. However not all of them have been measured across the studies which are included in our data catalogue. The charts below **(Figure 4-5)** display the number of studies in which not only these variables but also some other important variables have been observed.

**Figure 4.**
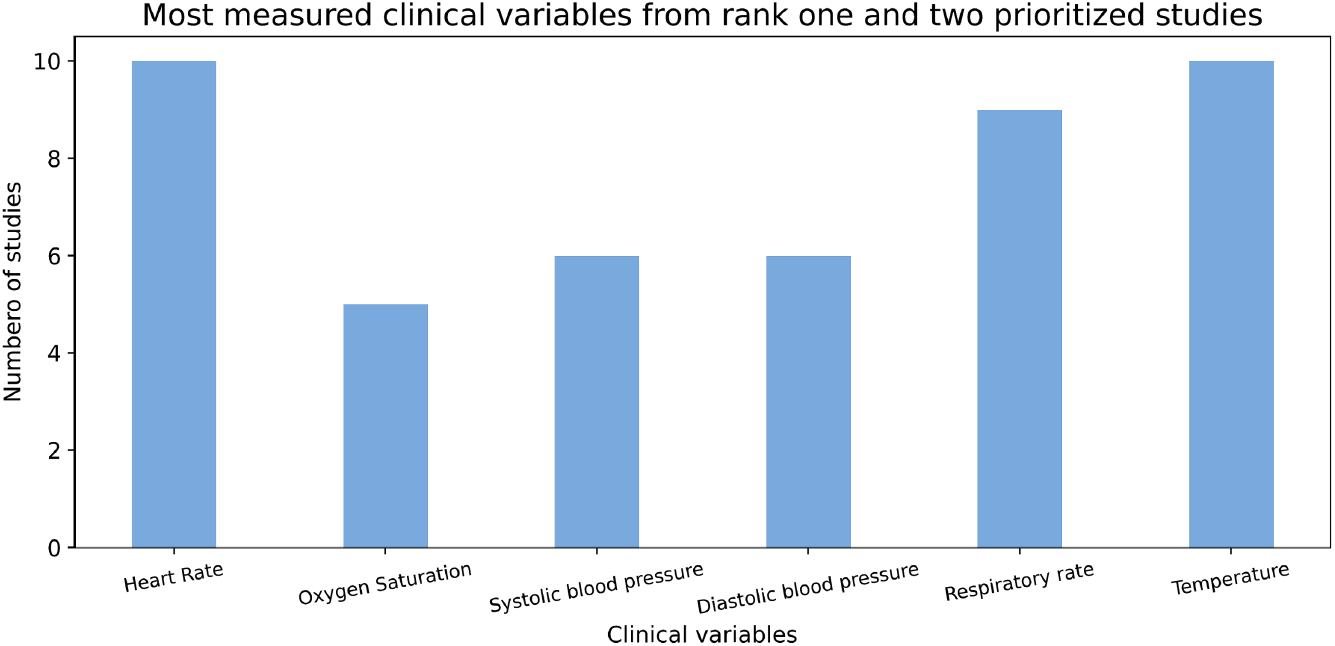
Distribution of different clinical variables from over 50 rank one and two prioritized studies. The X-axis shows the more common clinical variables, and the Y-axis shows the number of studies that these variables have been measured in. Heart rate and body temperature are the two most measured features from over 150 studies, with measurements observed in 10 collected studies.

**Figure 5.**
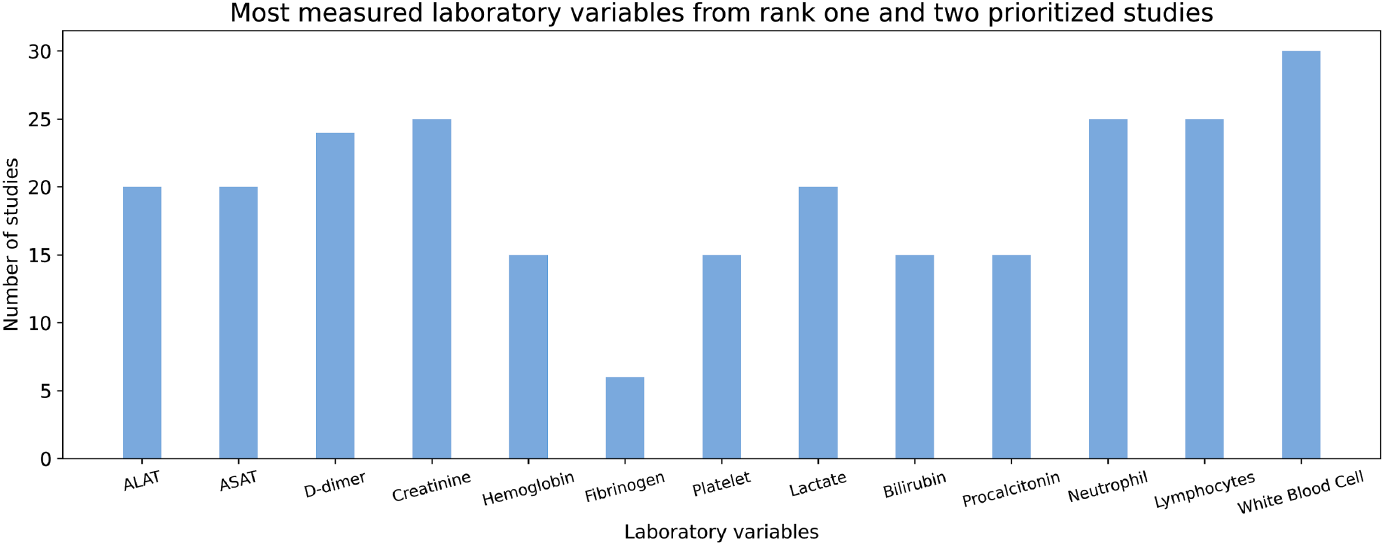
Distribution of different laboratory variables from over 50 rank one and two prioritized studies. The X-axis shows common laboratory variables while the Y-axis shows the number of studies that each of the variables have been measured in.

### 3.3. Data acquisition outcome

Study contacts (PIs) of collected studies were contacted in two rounds. In the first round, a data request email was sent (organized via a professional CRM system) to 552 contacts, of which 319 were from China. As we received a limited response from the contacts of each of these studies, we performed a second round of email-campaigning to 225 PIs who had not responded in the first round. In a third attempt, study contacts from 50 studies from rank one and two studies based on specific study regions (i.e., USA and European countries) and number of participants (i.e., more than 100) were prioritized and contacted directly via email. Overall, from the 552 data collection requests that were sent, only 15 replies were received, and the acquisition of exclusively four datasets was successful.

### 3.4. Summary of harmonization and variable mapping

While a wide range of studies have been performed since the start of the pandemic, not all of them have the same goal and study design and thus, their measured variables are different. For example, while in LEOSS and Medical Information Mart for Intensive Care (MIMIC) **[22]** studies, a wide range of features such as clinical, ICU, laboratory, drug and comorbidities have been measured, in other studies, such as data from COVID-19 patients from the Aachen and Frankfurt university clinics, ICU and laboratory features are the main measured features. To better assess the interoperability of the collected studies, we harmonized and mapped the corresponding variables of the studies to GECCO, COVOC and the COVID-19 ontology (see section 2.4.). **Table 2** represents the overview of interoperability evaluation by summarizing the numbers of features in individual studies and the number of common concepts between certain pairs of studies.

**Table 2.** Overview of the numbers of concepts in individual studies and the number of common concepts between pairs of studies.

| Dataset | Total Number of Concepts | Dataset | Total Number of Concepts | Common Concepts |
| --- | --- | --- | --- | --- |
| GECCO | 967 | LEOSS | 2,125 | 313 |
| GECCO | 967 | Aachen | 96 | 25 |
| GECCO | 967 | COVID-19 Ontology | 2,268 | 162 |
| GECCO | 967 | Coronavirus Vocabulary | 455 | 76 |
| GECCO | 967 | Erlangen | 530 | 18 |
| GECCO | 967 | Frankfurt | 74 | 4 |
| GECCO | 967 | MIMIC | 10,981 | 212 |
| GECCO | 967 | MC-19 | 106 | 24 |
| GECCO | 976 | Aachen | 96 | 25 |
| LEOSS | 2,125 | Aachen | 96 | 0 |
| Erlangen | 530 | Aachen | 96 | 16 |
| Frankfurt | 74 | Aachen | 96 | 8 |
| Erlangen | 530 | Aachen | 96 | 16 |

### 3.5. Dataset preparation for machine learning pipelines

As a showcase, **Table 3** summarizes the AI-readiness evaluation for one of the acquired datasets, Major Determinants of COVID-19 Associated Pneumonia study (MC-19), by investigating some of its main characteristics **(Figure 6-9)**. The procedure for the preprocessing of the dataset from this study is provided in the **Supplementary File**.

**Table 3.** AI-readiness evaluation table for the MC-19 dataset.

|  | Criteria | Quality |  |  |
| --- | --- | --- | --- | --- |
|  |  | Low | Medium | High |
| MC-19 | Number of patients | x |  |  |
|  | Completeness of data | x |  |  |
|  | Number of features in relation to number of patients |  | x |  |
|  | Event-rate (i.e, the fraction of patients with complete longitudinal follow-up and thus recorded events) |  |  | x |
|  | Objectiveness of the clinical outcome (i.e., is clinical outcome a subjective or physician-dependent variation?) | x |  |  |

**Figure 6.**
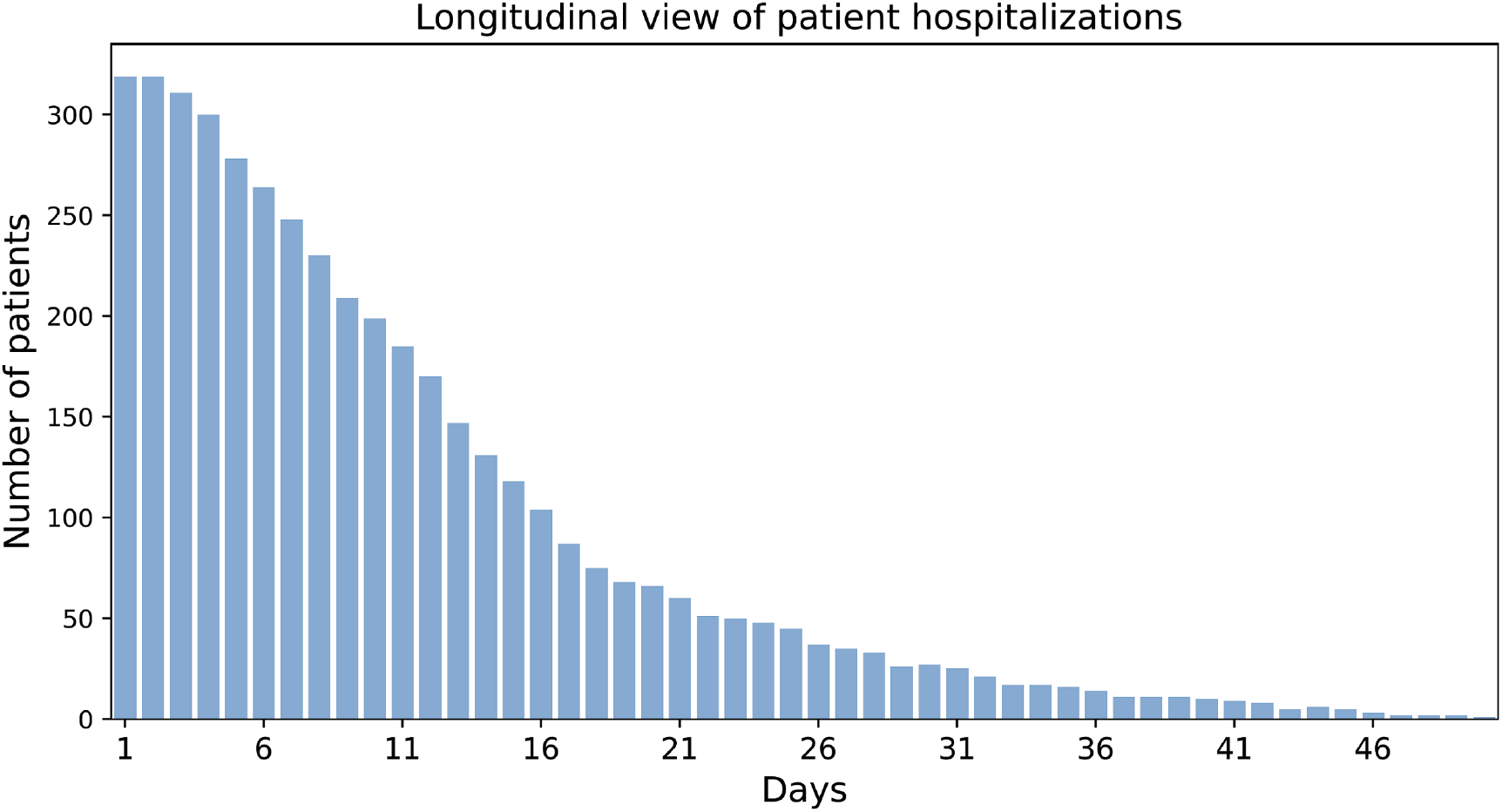
Longitudinal view of patient hospitalizations. The chart illustrates the duration of hospitalization (in days) for patients of the MC-19 study and thereby, inherently patient hospitalization outcomes (i.e., discharge, die). For most patients, hospitalization time was less than 14 days.

MC-19, sponsored by the Catholic University of the Sacred Heart in Italy, is an observational study (case - control) to assess if samples with negative tests in laboratory testing for COVID-19 suffering from pneumonia are positive in the serology for SARS-CoV-2. Moreover, the investigators aimed to identify patients with initially negative laboratory tests for COVID-19 with the help of serology measurements and CT scans.

#### Sample size and event rate

We visualized the longitudinal hospitalization of the patients and thus, by extension, the patient hospitalization outcome (i.e., discharged or died) over the course of hospitalization. While the hospitalization time was less than 14 days for most patients, some stayed up to 50 days in hospital **(Figure 6)**.

While MC-19 has 319 samples, hospitalization outcome was only recorded for 304 samples (i.e., event rate = 95%) and thus, the further downstream data-driven modelling would be limited to 304 samples. Although meaningful findings could be extracted from a dataset with only hundreds of samples, due to the limited sample size, the findings may not be reliable and affect the model generalizability. For example, investigating MC-19 shows the high risk of fatality in samples with body mass index (BMI) lower than 20 and higher than 30 (**Figure 7)**, which also has been shown in other studies **[23]**. However, due to the limited number of patients with the aforementioned BMI in MC-19, this finding can not be generalized.

**Figure 7.**
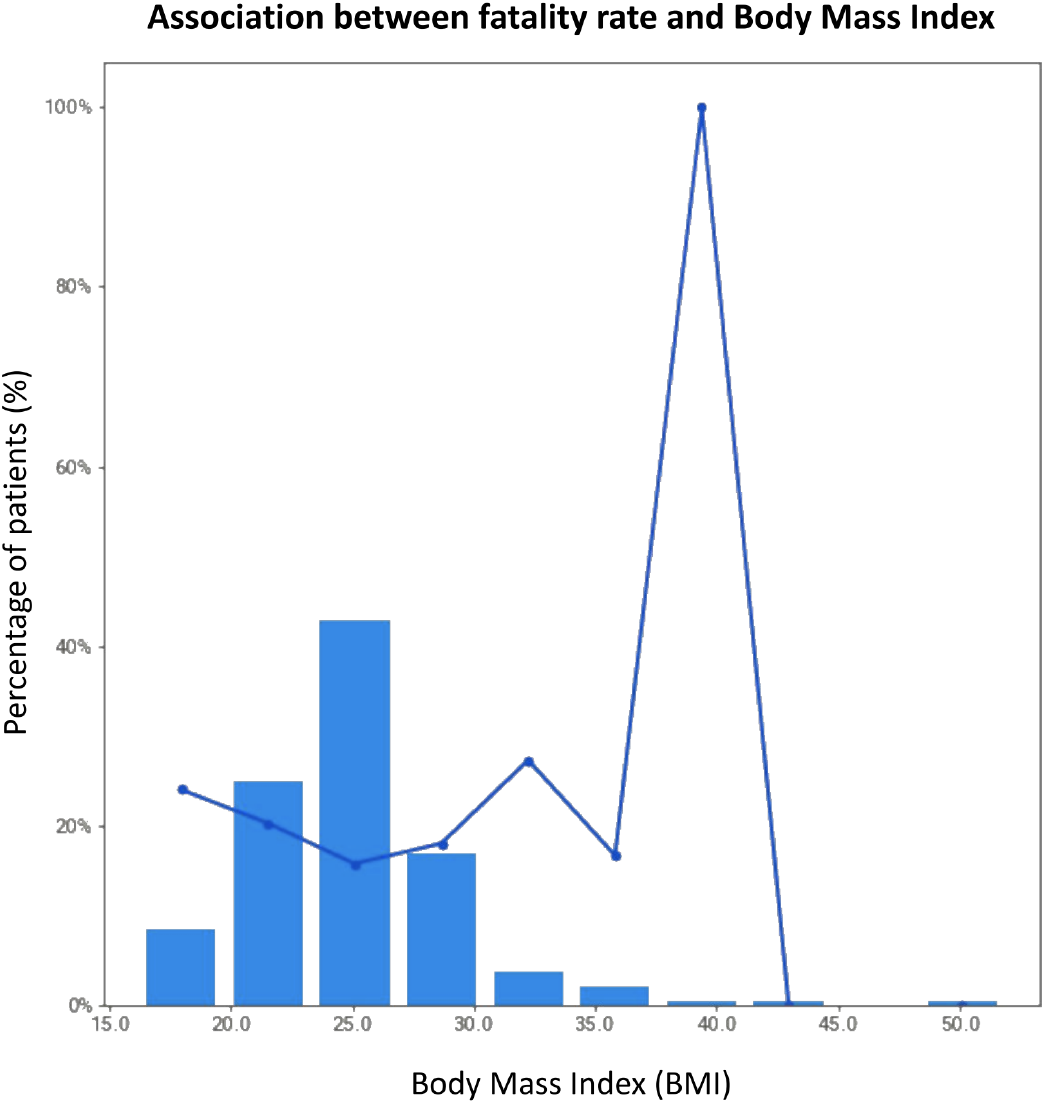
Association between fatality rate and BMI. The bars depict the percentage of patients for a range of BMIs, while the line graph displays information on the fatality rate for these patients. The fatality rate for patients with a BMI less than 20 or greater than 30 is highest among all other BMI ranges.

#### Data completeness

A wide range of variables, including drug, demographics, laboratory, serology, imaging, ICU and clinical variables were measured for this study **(Figure 8-9)**. We investigated the proportion of missing values for some of the longitudinal **(Figure 8)** and static variables **(Figure 9)**. While the proportion of missing values in static variables except “smoking” and “pneumonia” is not considerable, the proportion for longitudinal variables is significant. For example, measurements for “hemoglobin”, “hematocrit” and “Sio2” were not taken in half of the patient’s hospitalization time in almost 50% of patients. This much missing data could make a dataset unsuitable for applying statistics and further disease modelling.

**Figure 8.**
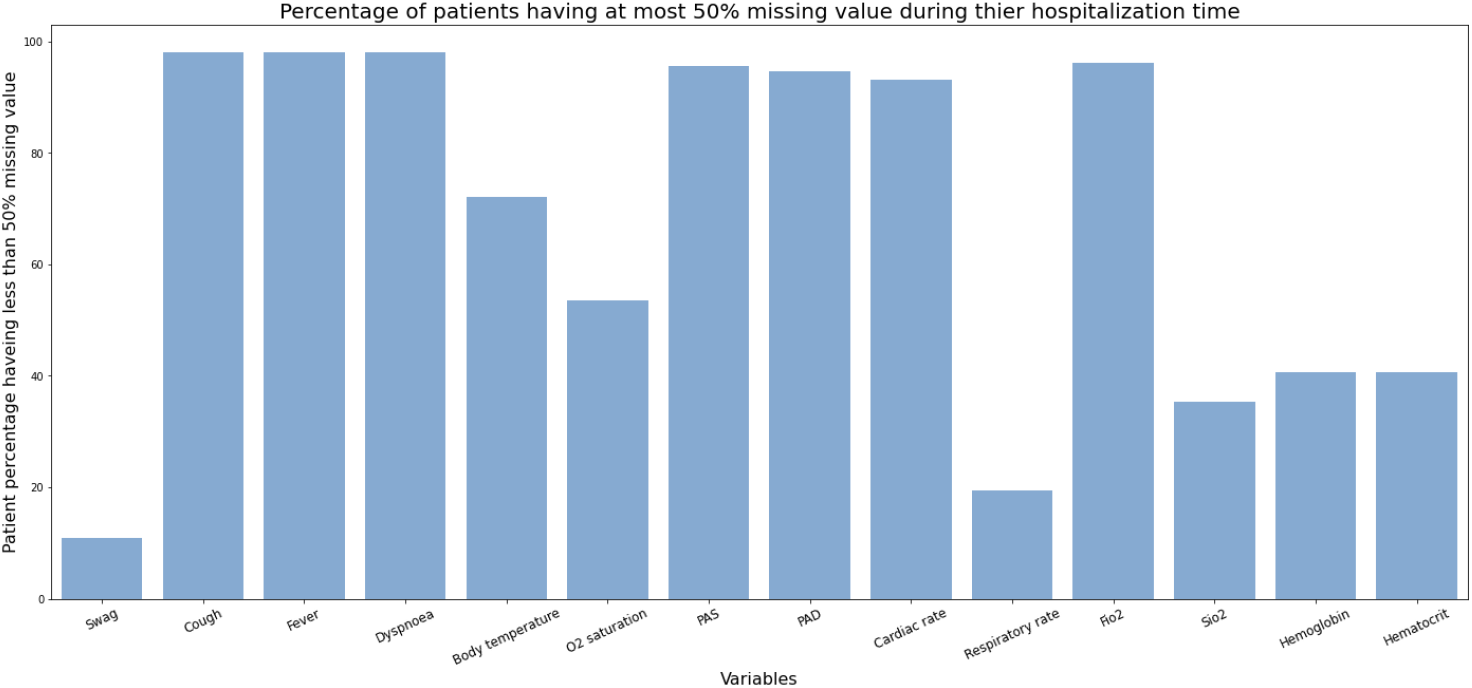
Percentage of patients having at most 50% missing values during hospitalization time. Hemoglobin, hematocrit and Sio2 have not been measured in half of the patient’s hospitalization time in almost 50% of patients. However, PAS, PAD, and Fio2 have been measured for more than half of the patient’s hospitalization time in almost 95% of patients.

**Figure 9.**
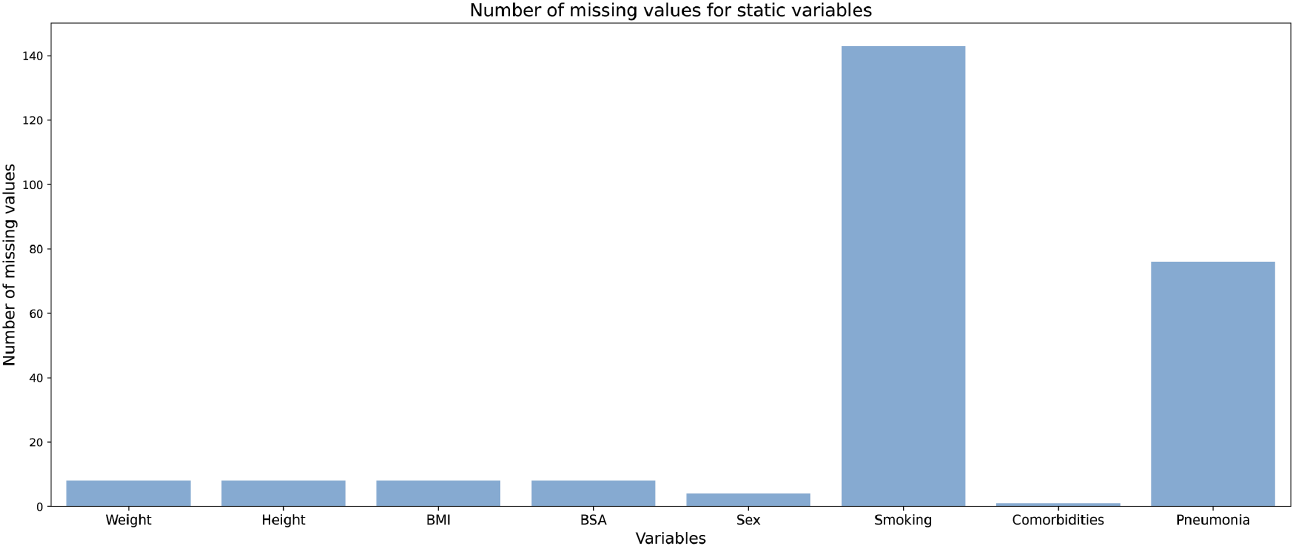
Number of missing values for static variables. The X-axis shows static variables, and the Y-axis shows the number of missing values for each variable. Smoking and pneumonia status are missing for more than 30% of patients.

#### First effort to aggregate cohorts

As the final target for making datasets ready for advanced algorithms is the aggregation of several cohorts to generate a consistent but expanded dataset for modelling applications, we attempted to integrate two of the sampled cohorts (MC_19 and Presurv**[24]**) into a new one (Merged). This effort required the fulfillment of several criteria to be technically and ethically successful. Firstly, only “matchable” variables were mapped and considered for aggregation (38 variables); secondly, patient identifiers and all related demographic variables remained unchanged during the merging process. Thirdly, visit dates were altered, removing their time notations and transforming them to an additional variable, “hospitalization days’’ (i.e., days spent in hospital since initial hospitalization). Finally, we annotated the provenance to maintain a track of the original entries. With these basic principles in mind, we generated the Merged cohorts. Our expectation of the Merged cohort may sound counterintuitive: we could not expect, for instance, a reduction in the absolute number of missing values, though we did expect a more coherent and less dispersed distribution of variable values, which we indeed found, as summarized in **Table 4**. Indeed, measuring the Absolute Distribution around Median (MAD), we found that the Merged cohort was more coherent and with fewer outliers than the individual, original cohorts for more than 58% of the aggregated variables. Naturally, for half of the variables, Merged showed a lower MAD value than MC_19, while for the other half of the variables, Merged was more coherent with respect to the Presurv cohort. These values were somewhat surprising as, while we could anticipate that variable ranges would be less dispersed in the Merged cohort with respect to the individual, original ones, we did not expect that they would be to the degree we observed. How much this behaviour is general and extendable to all cohorts is beyond the scope of the present paper and we will perform a deeper analysis on this in future.

**Table 4.** Summary of MAD values for matched numeric variables of differing provenance. The table reports a set of 22 variables present in each of the cohorts (58% of 38 toal numeric variables). Lower MAD deviations from the median signifies that cohort aggregation helped make variables more coherent or closer to the median.

| Sr. | Term | MAD (Median Absolute Deviations) Statistics |  |  |
| --- | --- | --- | --- | --- |
|  |  | MC_19 | Merged | Presurv |
| 1 | Hospitalization days | 10.4 | 10.4 | 3.0 |
| 2 | Eosinophils count | 0.1 | 0.1 | 0.0 |
| 3 | Total proteins | 8.9 | 7.4 | 6.8 |
| 4 | Procalcitonin | 0.2 | 0.1 | 0.1 |
| 5 | Alkaline Phosphatases | 28.6 | 28.2 | 26.7 |
| 6 | Hematocrit | 6.1 | 5.5 | 4.7 |
| 7 | Eosinophils percentage | 1.2 | 0.9 | 0.1 |
| 8 | Monocyte count | 0.2 | 0.2 | 0.2 |
| 9 | Sodium | 4.4 | 4.3 | 4.2 |
| 10 | Platelets count | 123.1 | 118.6 | 101.6 |
| 11 | Gamma glutamyl transpeptidase | 32.6 | 26.7 | 22.2 |
| 12 | Interleukin 6 | 35.6 | 28.7 | 25.8 |
| 13 | Neutrophil count | 3.7 | 4.3 | 4.7 |
| 14 | Monocytes percentage | 2.2 | 2.8 | 4.3 |
| 15 | Ferritin | 570.1 | 678.9 | 706.0 |
| 16 | Neutrophils percentage | 12.6 | 15.9 | 17.6 |
| 17 | Lymphocytes percentage | 10.1 | 12.0 | 12.9 |
| 18 | Potassium | 0.6 | 0.6 | 0.7 |
| 19 | pH | 0.0 | 0.1 | 0.7 |
| 20 | WBC | 4.2 | 4.6 | 4.7 |
| 21 | RBC | 0.7 | 0.7 | 0.7 |
| 22 | HCO<sub>3</sub> | 3.0 | 3.4 | 3.7 |

### 3.6. COVID-19 Datasets Visualization

The COVID-19 data viewer (https://www.coperimoplus.eu/en/data-models/data.html) summarizes the most important metadata, such as measured features, facilitates accessing the datasets and visualizing the relative number of collected studies in the data catalogue based on their geographic location **(Figure 10)**. Based on the geographic distribution of studies, the number of collected studies from European countries, such as Sweden and Russia, as well as countries from South America, such as Brazil, are greater compared to other locations. Furthermore, we released the underlying data at https://github.com/SCAI-BIO/covid-data-catalogue.

**Figure 10.**
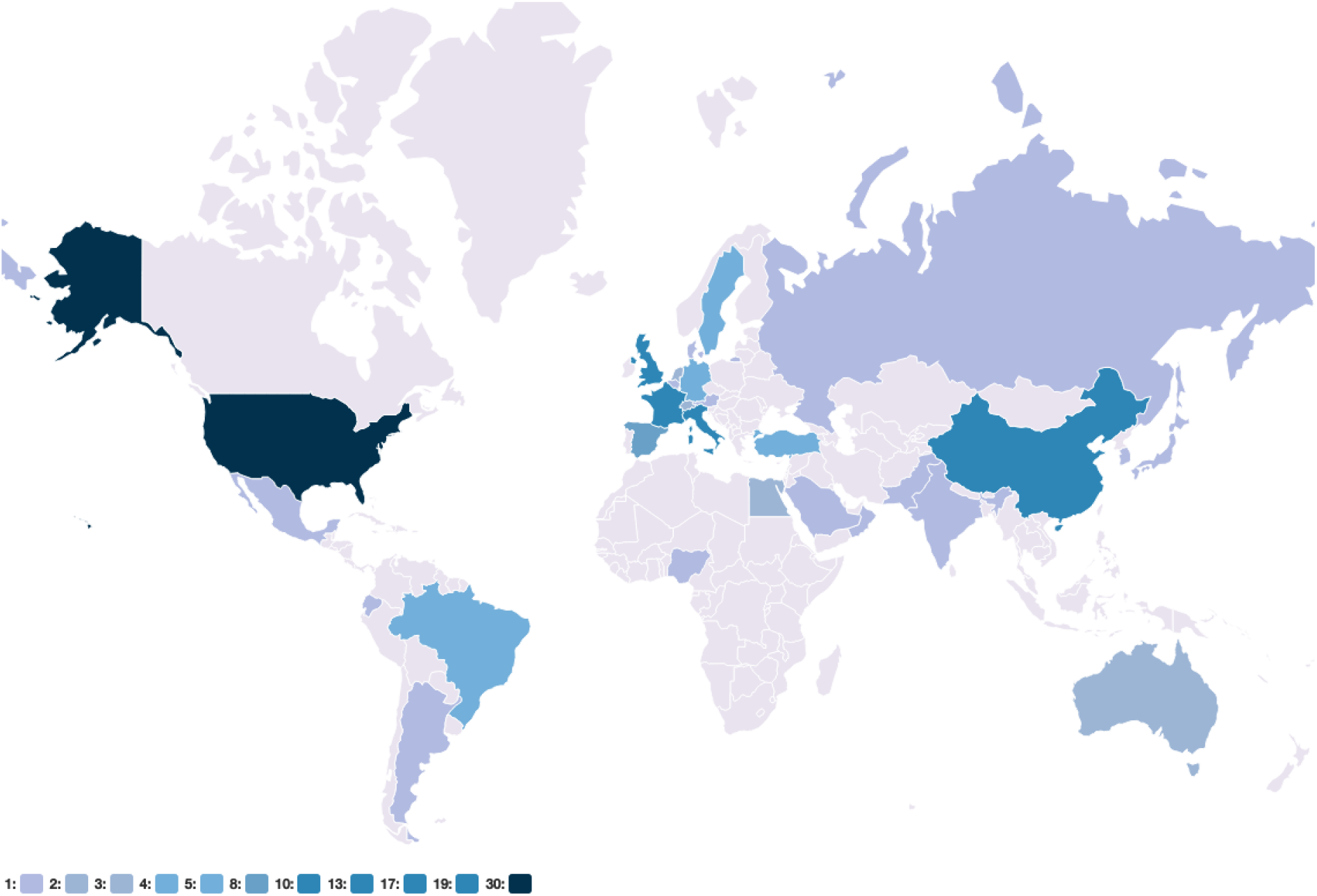
Geographic distribution of studies. The relative number of studies in each country is highlighted in blue. The underlying data is available at https://github.com/SCAI-BIO/covid-data-catalogue/blob/main/data/general.tsv.

## 4. Discussion

Our data catalogue provides information on 553 studies on COVID-19, identified from manuscripts, clinical trials, and global initiatives. The organization of these studies supports the development of personalized predictive models and the broad investigation of disease risk factors which, in turn, may lead to opportunities for therapeutic interventions. The data catalogue involved significant curation, harmonization and evaluation of datasets which were not free of challenges. The challenges are discussed in three main groups including sociological and technical and AI-readiness evaluation.

### Ethical and sociological challenges

While we submitted data access requests to more than 500 PIs, only a handful of them shared their data. There were multiple reasons due to which we could not get access to some of the cohorts. First, the majority of data owners did not reply to our inquiries despite sending multiple follow-up emails over the course of several months and even despite attempts to connect via business networks and social media. Second, legal issues during the data sharing process hindered our access to datasets. Third, ethical considerations also hampered access to datasets. Participant data can potentially be associated with their identity and, accordingly, data sharing poses a risks of identification of participants as well as stigmatization and discrimination which ultimately prevents data owners from sharing their data. Hence, privacy interests of participants are also important considerations to be made.

### Technical challenges

The lack of interoperability across datasets impedes the development of harmonized datasets as each of the datasets were established without the use of controlled vocabularies for consistent feature naming. Thus, even if the same feature was collected in two studies, it would be referred to in different terms, severely impeding the direct comparison of datasets and validation of findings across disparate cohorts.

### AI-readiness evaluation

The readiness of a dataset for AI purposes can be evaluated based on multiple criteria, each of which are highly situation dependent. For example, while hundreds of samples can lead to a good performance using a simple linear AI algorithm aimed at predicting the true relationship between patient-specific characteristics and the endpoint, a non-linear AI algorithm such as neural networks may require thousands of samples. Moreover, the number of samples in a dataset should be representative of the entire disease population. Thus, modelling approaches which are based on a few hundred samples might not represent the heterogeneity of the entire population. Given these two aforementioned points, while the MC-19 dataset can be used for simple time-to-event models, it might not be practical to be used with powerful AI algorithms for personalized predictive models as it has a limited number of samples. In addition, it is important to consider the completeness of the data as it is generally consistent over all features in a dataset. Thus, removing missing values, imputing missing values, and removing features with a high number of missing values would be three potential solutions. While the first solution (i.e., removing missing values) may influence the number of features in relation to the number of patient criteria, the second option (i.e., imputation) could lead to model inaccuracy and affect the model robustness. Removing the features with a high proportion of missing values could also lead to losing a lot of information, which in turn causes model unreliability. The applicability of MC-19 with approximately 50% missing values specifically for variables with follow-up measurements highly depends on the type of AI algorithm. Taken together, for a dataset to be “AI ready”, which might need a lot of preprocessing work, is highly subjective and different factors should be taken into consideration.

In summary, the presented data catalogue organizes knowledge in the field of COVID-19 and provides a comprehensive overview of hundreds of studies, bringing together data from various studies so that researchers can understand what has been measured, in which population and under what clinical context. Finally, the catalogue can be used for data-driven COVID-19 model development, such as personalized predictive models. This first version of the COVID-19 data catalogue involved significant curation and harmonization of information from disparate studies. The data catalogue is complemented with a comprehensive web application that provides interactive visualizations and tools to query the catalogued knowledge.

As this resource expands to capture all known data around COVID-19, we aim to facilitate dataset interoperability by developing a comprehensive, COVID-19 common data model, which could support the alignment and mapping of variables by providing easy-to-follow guidelines and a dedicated interface for retrospective data harmonization. This in turn would facilitate more formal meta-analyses so that robust conclusions may be drawn around the state of the field, in turn leading to new hypotheses for future studies.

## Data Availability

The data should be applied from Catholic University of the Sacred Heart

## Acknowledgement

This research was conducted in the context of the ‘COPERIMO*plus*’ initiative and supported by the Fraunhofer ‘Internal Programs Fraunhofer vs Corona’ under Grant No. Anti-Corona 840266.

## Footnotes

1 https://www.worldometers.info/coronavirus/

