## Supplementary files for "Curating, collecting, and cataloguing global COVID-19 datasets for the aim of predicting personalized risk"

### Supplementary File

#### Dataset preparation for AI-readiness evaluation

**Data cleaning:** Duplicates were removed. Missing values were encoded consistently to avoid ambiguity. Special characters were removed or replaced if applicable. Ambiguous feature representations were replaced with unique representations or unique identifiers if applicable, e.g. mapping drug brand names to substance name. We checked feature distributions for potential outliers and removed or corrected questionable values if applicable. In addition, we performed random spot-checks on observations.

**Data transformation:** The data was restructured to provide one table per modality (e.g., diagnosis, treatments, lab, demographics) and clinical outcome variables. Longitudinal observations were separated from the non-longitudinal ones.

**Metadata provision:** A data dictionary was compiled, containing variable names, descriptions, data types, value ranges for numerical variables, encodings and decodings for categorical variables and units of measure.

#### Examples of relevant and non relevant trials from the Chinese Clinical Trial Registry (ChiCTR)

As an example, selected studies include, highly relevant: ChiCTR2000029757 (with highly relevant outcomes including ICU related data), ChiCTR2000030254 (with highly relevant outcomes including ICU related data); relevant: ChiCTR2000029765 (with relevant outcomes (e.g., mortality rate)); and not relevant: ChiCTR2000030046 (has only 10 participants), ChiCTR2000029770 (with sequential study type).

**Supplementary Table:**

| Class | Outcome measures |
|---|---|
| **Highly relevant** | Mortality outcome in ICU patients, Oxygen saturation, Intubation, In-hospital mortality, Length of hospitalization, Efficacy of CT scan and serology, Infection severity, causes of ICU admission, ICU survival, Duration of mechanical ventilation, Complications during the ICU stay, Characteristics of pulmonary CT-scan, All-cause death, ICU hospitalization,Ventilation analysis, Incidence of thromboembolic events, ICU-free days alive from ICU-admission, Degree of severity of respiratory disease, Septic shock as defined by sepsis-3 criteria, Sepsis-induced coagulopathy (SIC) , Rapid Severity Index for COVID-19 (qCSI) |
| **Relevant** | Prognostic factors of severe state, Clinical and radiological characteristics, Biological parameters, Physical examination data, Seroprevalence (Hospital in a Low Incidence Area), Posteroanterior identification of COVID-19 (Hospital in a Low Incidence Area), Anti-infective agents impact, Possible Predictors of Mortality, Possible Predictors of Mortality, Infection Associated With Endoscopy, Predictive performance (based on Clinical and Radiomic Model), Mortality outcome (ML approach) |
| **Less relevant** | Mortality (appendicitis patients), Postoperative length of stay (appendicitis patients), Factors affecting prognosis (pregnant women), D-dimer values (pregnant women), Blood count parameters (pregnant women), Covid attack rate (HIV and/or on PrEP patients), Risk factors (Kidney Transplant Recipients), Antibody positivity (health care workers), Rate of Death (patients with acute kidney injury), Length of hospital stay (patients with acute kidney injury), Mortality (cancer patients), Hospitalizations (cancer patients), Hospitalizations (cancer patients), Overall survival (cancer patients) |

**Supplementary Table 1:** The complete list of criteria and different outcome measures considered to classify the clinical trials.
